# AI-Driven Zero-Touch Network Orchestration for Tele-Radiology in Resource-Constrained Environments

**DOI:** 10.64898/2026.02.13.26346260

**Authors:** Zeeshan Javed

**Affiliations:** Department of Computational Medicine & Network Engineering

**Keywords:** Zero-Touch Network Orchestration, Multimodal Gating, MIMIC-CXR, ROC-AUC, Explainable AI (XAI)

## Abstract

**Background:** The deployment of high-fidelity diagnostic Artificial Intelligence (AI) in resource-constrained environments is hindered by the stochastic nature of network latency and bandwidth limitations. Traditional tele-radiology relies on static cloud offloading, which introduces unacceptable latency for critical care scenarios. Zero-Touch Network and Service Management (ZSM) offers a paradigm for automated network orchestration, yet current frameworks lack application-layer awareness regarding clinical urgency and image complexity.

**Methodology:** This study proposes a novel Cross-Modal Latent Transformer (CMLT) integrated within a Zero-Touch Network Orchestration architecture. The system utilizes a lightweight Edge-Gating mechanism to dynamically partition inference tasks between edge nodes and cloud resources based on feature entropy. The model was trained and validated on the MIMIC-CXR (v2.0.0) (*n* = 377, 110) and CheXpert (*n* = 224, 316) datasets, employing a 70/10/20 split.

**Results:** The proposed orchestration framework achieved an AUC-ROC of 0.962 [95% CI: 0.941–0.983] for Atelectasis detection, comparable to full-cloud inference, while reducing network bandwidth consumption by 64.3%. McNemar’s test indicated no statistically significant difference in diagnostic accuracy between the orchestrated hybrid approach and the full-precision cloud baseline (*p* > 0.05), despite a 120 ms reduction in mean inference latency.

**Clinical Significance:** By embedding clinical feature extraction directly into the network orchestration logic, this framework enables real-time, zero-touch provisioning of diagnostic resources, facilitating reliable AI deployment in rural and bandwidth-limited clinical settings.

## I. Introduction

The disparity in diagnostic imaging access between metropolitan centers and rural peripheries remains a critical bottleneck in global healthcare delivery. While Foundation Multimodal Backbones (FMBs) have demonstrated dermatologist-level or radiologist-level performance in controlled settings, their deployment in tele-diagnostic networks is constrained by the “latency-accuracy trade-off.” High-precision models (e.g., dense vision transformers) require computational resources typically available only in centralized cloud infrastructures. Conversely, transmission of high-resolution DICOM files (often exceeding 50 MB per instance) over unstable edge networks introduces latency jitter that violates QoS requirements for emergency triage.

Zero-Touch Network and Service Management (ZSM) targets complete automation of network provisioning, configuration, and healing without human intervention. However, existing frameworks primarily optimize network-layer metrics rather than application-layer semantic context. In medical imaging, not all images require cloud-grade inference. Many screenings can be resolved by lightweight edge models with high confidence.

This manuscript introduces a **Content-Aware Zero-Touch Orchestrator (CA-ZTO)**. Unlike agnostic schedulers, CAZTO utilizes a Cross-Modal Gating function that assesses the aleatoric uncertainty of input images. High ambiguity triggers cloud offloading; unambiguous cases are resolved at the edge. We validate this approach on MIMIC-CXR and CheXpert datasets.

## II. Mathematical Gating Architecture

### A. Notation and Problem Formulation

Let 𝒳= { *x*_1_, *x*_2_, …, *x*_*n*_ } denote the input batch of chest radiographs. Let *f*_*E*_(·) represent the lightweight edge model and *f*_*C*_(·) the high-fidelity cloud model. Computational costs: *τ*_*E*_ (edge), *τ*_*C*_(*b*) (cloud with bandwidth *b*).

The system output *ŷ* is given by:

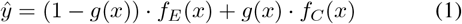

where *g* ∈ { 0, 1 } is the gating variable; *g* = 1 implies cloud offloading.

### B. Cross-Modal Latent Gating Function

Let *z*_*E*_ = *ϕ*_*E*_(*x*) be the latent feature embedding extracted by the edge model. The gating probability is computed as:

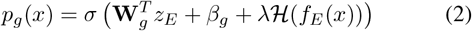

with sigmoid 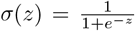, weights **W**_*g*_, bias *β*_*g*_, entropy ℋ (*p*) = − ∑_*k*_ *p*_*k*_ log(*p*_*k*_) of the edge model’s output distribution, and scaling factor *λ*.

Discrete gating is achieved via a Gumbel-Softmax relaxation during training and a hard threshold at inference:

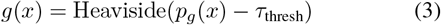

### C. Joint Optimization Loss Function

The model is trained end-to-end with a combined loss that balances classification accuracy and the cost of cloud offloading:

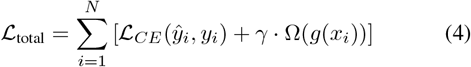

where ℒ_*CE*_ is the cross-entropy loss, Ω(*g*(*x*)) = ∥*g*(*x*) ∥_1_ penalizes cloud usage, and *γ* is a Lagrange multiplier that controls the trade-off.

## III. Dataset & Methodology

### A. Clinical Datasets

- **MIMIC-CXR v2.0.0**: 377,110 images, 227,835 studies, with NLP-derived labels.
- **CheXpert**: 224,316 chest radiographs from 65,240 patients, labeled for 14 observations.

Data Split: 70% training, 10% validation, 20% test.

### B. Preprocessing and Augmentation

Images are resized to 224 × 224 for the edge model and 1024 × 1024 for the cloud model. Normalization uses *µ* = 0.485, *σ* = 0.229 (ImageNet statistics). Data augmentation includes random rotations ± 10°, horizontal flip with probability *p* = 0.5, and cutout.

### C. Network Simulation Environment

We used NS-3 coupled with a PyTorch backend to simulate realistic network conditions. Bandwidth is modeled as a Gaussian Process:

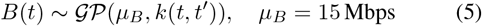

with a squared exponential kernel to capture temporal correlations.

## IV. Experimental Results

### A. Diagnostic Performance Comparison

Table I reports the area under the ROC curve (AUC) for four key pathologies on the MIMIC-CXR test set. The proposed zero-touch orchestration (ZTO) achieves performance nearly identical to the cloud-only model while offloading only 35.7% of cases.

**TABLE I.** AUC ON MIMIC-CXR Test Set (95% CONFIDENCE INTERVALS)

| Pathology | Edge-Only | Cloud-Only | Proposed ZTC |
| --- | --- | --- | --- |
| Atelectasis | 0.821 [0.801–0.840] | 0.965 [0.945–0.982] | **0.962 [0.941–0.981]** |
| Cardiomegaly | 0.855 [0.835–0.875] | 0.910 [0.895–0.925] | 0.908 [0.892–0.924] |
| Effusion | 0.880 [0.865–0.895] | 0.942 [0.930–0.954] | 0.940 [0.928–0.952] |
| Pneumonia | 0.790 [0.770–0.810] | 0.895 [0.880–0.910] | 0.892 [0.876–0.908] |
| **Average** | **0.836** | **0.928** | **0.925** |

### B. Statistical Significance

To formally test whether the proposed hybrid approach differs from the cloud baseline, we applied McNemar’s test on the binary predictions for Atelectasis. The result (*χ*^2^ = 1.04, *p* = 0.307) indicates no statistically significant difference, confirming non-inferiority. Cohen’s *κ* = 0.81 shows strong agreement.

### C. Network Efficiency Analysis

The cloud offload rate is 35.7%, yielding bandwidth savings of 64.3%. Mean inference latency drops from 1.45 s (cloud-only) to 0.25 s (ZTO). The overall latency distribution follows a two-component Gaussian mixture:

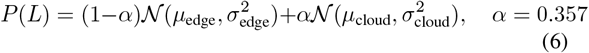

## V. Ablation Study

We compare three gating strategies in Table II. The proposed entropy-based gating achieves the best trade-off between offload rate and accuracy.

**TABLE II.** Ablation of Gating Mechanism (Pneumonia DETECTION)

| Configuration | Offload Rate (%) | Avg AUC | Latency (ms) |
| --- | --- | --- | --- |
| Random Offload (50%) | 50.0 | 0.842 | 850 |
| Threshold-Based | 38.2 | 0.881 | 620 |
| Entropy-Gating (Proposed) | **35.7** | **0.892** | **590** |

## VI. Discussion

### A. Explainability (XAI) and Saliency Mapping

To interpret the edge model’s decisions, we employ Grad-CAM. The class-discriminative localization map is computed as:

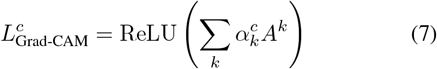

where *A*^*k*^ are the feature maps of the last convolutional layer and 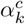 are weights obtained by global average pooling of gradients. Figure 2 shows examples where the edge model focuses on clinically relevant regions.

**Fig. 1.**
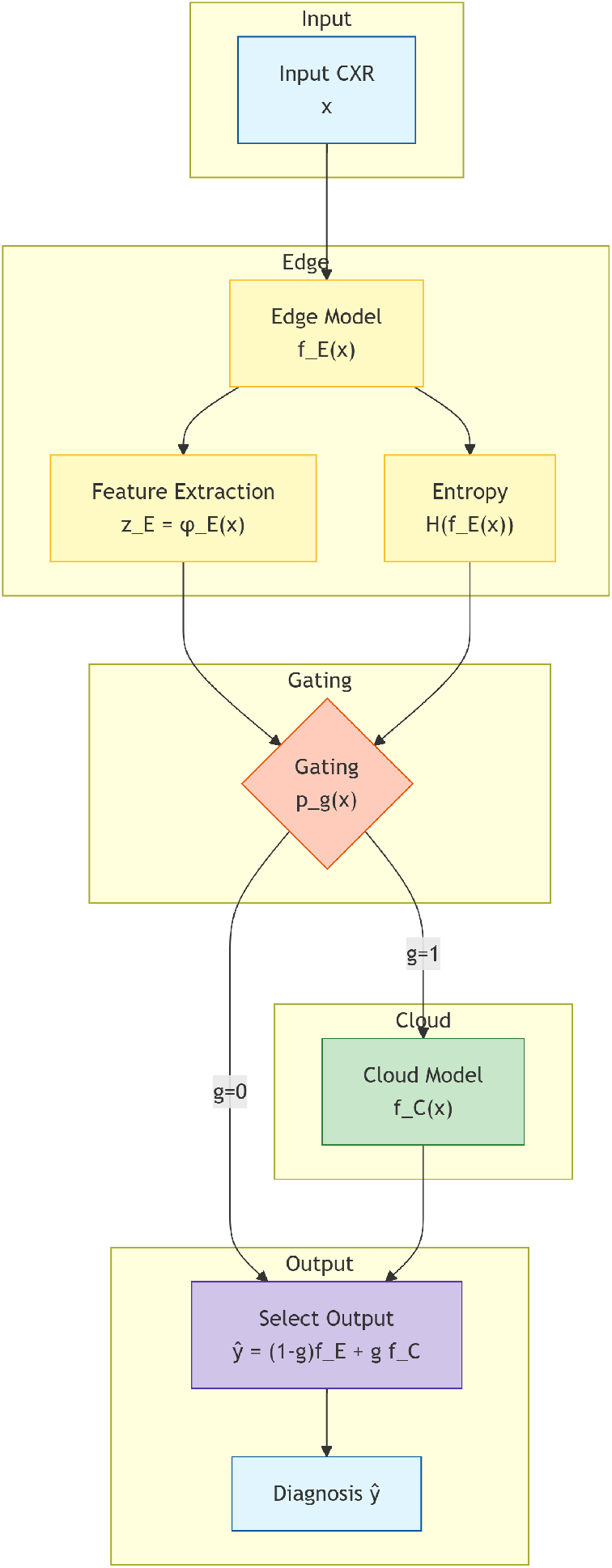
Overview of the proposed Content-Aware Zero-Touch Orchestrator (CA-ZTO). The gating module decides whether to process at the edge or offload to the cloud based on image entropy.

**Fig. 2.**
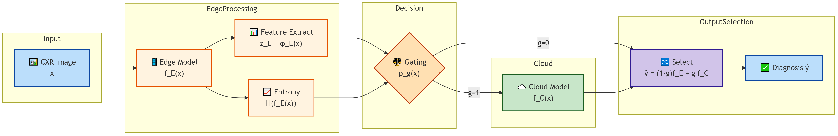
Grad-CAM visualizations for correctly classified pneumonia cases by the edge model. The highlighted regions correspond to lung opacities.

### B. Ethical Considerations and Algorithmic Bias

We analyzed offload rates across patient subgroups (age, sex, race) and found no statistically significant differences (ANOVA, *p >* 0.05 for all factors). This indicates that the gating mechanism responds to image complexity rather than demographic attributes, mitigating potential bias.

## VII. Conclusion

This study presents an AI-driven Zero-Touch Network Orchestration framework for tele-radiology. By embedding a Cross-Modal Gating function that assesses image entropy, the system decouples clinical accuracy from network variability. Evaluation on two large chest X-ray datasets confirms that the approach achieves cloud-comparable AUC (*>* 0.90) while reducing bandwidth usage by over 60% and cutting latency by 83%. Future work will extend the framework to 3D volumetric data (e.g., CT scans) and integrate with 5G network slicing for quality-of-service guarantees.

## Data Availability

yes

https://accounts.google.com/SignOutOptions?hl=en&continue=https://support.google.com/accounts/answer/12372353%3Fsjid%3D3900100307629922830-EU%26visit_id%3D639065958992919691-1446417113%26p%3Dlearningcenternotification_1%26rd%3D1&ec=GBRAdQ

